# Altered placental phenotype and increased risk of placental pathology in fetal spina bifida: a matched case-control study

**DOI:** 10.1101/2023.09.20.23295859

**Authors:** Marina White, David Grynspan, Jayden Arif-Pardy, Tim Van Mieghem, Kristin L Connor

## Abstract

Open spina bifida (SB) remains one of the most common congenital anomalies and associates with significant comorbidities in the fetus, which may, in part, be driven by placental maldevelopment. We hypothesised that placental pathologies and maldevelopment would be more prevalent in fetuses with SB compared to fetuses without congenital anomalies. Placental histopathology (H&E-stained slides) and transcriptome (Clariom D™ microarray) were evaluated for fetuses with isolated open SB undergoing either pregnancy terminations or term deliveries (cases; n=12) and control fetuses without congenital anomalies having term or preterm births (n=22). Associations between study group and placental histopathology were adjusted for fetal sex and gestational age [GA] at delivery. Relationships between placental histopathology and select placental gene expression signatures were also evaluated. Case placentae had lower placental weight than controls (median [IQR]: 263g [175, 370] vs. 455 [378, 560], p=0.001). Placental villi structural phenotype was different in cases, who had a higher proportion of immature intermediate villi than controls (32.5% [6.3, 56.3] vs. 10% [5, 13.8], p=0.01), but similar proportions of mature intermediate (10% [5, 10] vs. 10% [8.75, 11.25]) and terminal villi (22.5% [11.3, 43.8] vs. 30 [20, 36.3]), and similar odds of having many syncytial knots (adjusted odds ratio [aOR]=6 [0.2, 369]) as full-term born controls. Cellular phenotypic differences in case placentae included higher odds of having many Hofbauer cells (aOR=16.2 [1.4, 580], p=0.02) and a thick syncytial membrane (aOR=146 [3, 3.46e5], p=0.007). Expression in gene pathways related to immune/inflammatory processes, spinal cord injury, and Hedgehog and Wnt signaling were associated with placental maturity in cases. This study is the first to characterize placental histopathology in a contemporary cohort of fetuses with SB. Improved knowledge on placental histopathological and genetic phenotypes in spina bifida increases our understanding of mechanisms that may drive comorbidities to ultimately reduce offspring morbidity and mortality.

## Introduction

Spina bifida (SB), which accounts for approximately two-thirds of all neural tube defects (NTDs), affects around one per 1000 livebirths worldwide (1). SB associates with low birthweight (2, 3) and younger gestational age (GA) at birth (4), which increases the risk of mortality for affected children (5, 6) and may be linked to placental dysfunction (4). While both environmental and genetic factors are known to contribute to SB risk (7-9), only half of all SB cases are estimated to be responsive to current preventative strategies (10). Thus, there remains a need to expand knowledge on the causal mechanisms of SB and its comorbidities, to advance strategies for prevention and improve early developmental trajectories.

A suboptimal early pregnancy environment permissive of SB, which arises during the fourth week post-fertilisation, will also shape the development and function of early placental structures and may have lasting influence on placental function (11). Thus, placental maldevelopment in pregnancies with fetal SB may be a consequence of the same risk factors that led to SB, and this placental maldevelopment could contribute to the increased risk of poor fetal growth and development (12) and early birth (13) observed in fetuses with SB (2-4). However, placental development and function in pregnancies with fetal NTDs have not been well-studied. Understanding whether and how placental phenotype is altered in fetuses with NTDs could provide insight into mechanisms underlying NTD-associated comorbidities, and may even hold clues about the early pregnancy environment that resulted in improper neural tube closure.

We previously found that fetal NTDs associated with increased risk of placental pathologies in the Collaborative Perinatal Project cohort, which predated the introduction of mandatory folic acid fortification and the promotion of periconceptional folic acid supplementation in the United States (14, 15). Whether NTDs associate with increased risk of placental pathologies has not been investigated in a contemporary cohort.

Here we evaluated placental histopathology in placentae from fetuses with open SB and fetuses with no congenital anomalies, in a contemporary cohort from Toronto, Canada, where folate deficiency is rare (16). We hypothesised that cases would have increased risk of placental maldevelopment and pathologies compared to control fetuses. We also explored relationships between placental histopathology and placental transcriptome signatures, to determine whether expression of select gene networks associated with histopathological phenotype, and between placental histopathology and maternal and infant characteristics.

## Methods

### Ethics

The Carleton University Research Ethics Board (108884 and 106932) and the Mount Sinai Hospital Research Ethics Board (17-0028-E and 17-0186-E) approved this study. All procedures were performed in accordance with the Helsinki Declaration of 1975, as revised in 1983.

### Population and study design

This case-control study has been previously described in detail (17). In brief, we prospectively recruited patients carrying a fetus with open SB (cases) early in the third trimester of pregnancy through the Fetal Medicine Unit at Mount Sinai Hospital, Toronto. Patients carrying a fetus without congenital anomalies (SB controls) were recruited through our low-risk pregnancy unit around 25 weeks’ gestation. We also identified a second control group that was matched to cases for GA at delivery and maternal pre-pregnancy body mass index (BMI). These controls delivered preterm (PT controls) and had no other known pregnancy complications.

### Maternal clinical, demographic, and dietary characteristics

Maternal clinical data and medical history, and a 24-hour dietary recall (Automated Self-Administered 24-hour Recall™, specific to the Canadian population [ASA24-Canada-2016] (18)) were collected at recruitment for cases and SB controls. Methods used to analyse the maternal dietary recall data for this cohort have been previously described (17), and included calculations of estimated average requirements (EARs) (19), Healthy Eating Index (HEI)-2015 (20) scores, and dietary inflammatory index (DII) scores (21).

### Infant birth outcomes

Data on pregnancy outcomes and infant birth weight (adjusted for infant GA and sex using Canadian growth standards (22)) were collected at delivery.

### Placental collection and processing

Immediately after delivery, placental samples were collected and processed by the Research Centre for Women’s and Infants’ Health (RCWIH) BioBank. Using standard procedures, placental biopsies were collected from each of the four placental quadrants, at least 1.5 cm away from the centre and edge of the placental disk, formalin-fixed and paraffin embedded, and then sectioned (5 μm) and stained with haematoxylin (Gill No. 1, Sigma-Aldrich) and eosin (Eosin Y-Solution, Sigma-Aldrich; H&E), following standard protocols (23). Placental biopsies were also snap frozen in liquid nitrogen and stored at -80°C for whole transcriptome profiling (17).

### Placental phenotype

#### Anthropometry

Placental weight was recorded from fresh, untrimmed placentae prior to sampling. Raw measures of placental weight and GA-corrected placental weight z-scores were analysed (24). Ratios for infant birthweight-to-placental weight were used as an estimate of placental efficiency (25).

#### Histopathology

Placental histopathology was semi-quantitatively evaluated on H&E-stained slides by a clinical pathologist (DG), who was blinded to the study group, guided by the Amsterdam criteria (26). The primary outcomes were:

1. Characteristics of placental villous maldevelopment:
  a. delayed villous maturation (absent, present [≥10 monotonous immature intermediate villi with centrally placed capillaries and few vasculosyncytial membranes]);
  b. distal villous hypoplasia (absent, low grade [scarce and thin terminal and mature intermediate villi surrounding thin stem villi,], high grade [scarce and thin villi surrounding stem villi, syncytial knots present, in multiple focus areas]);
  c. accelerated villous maturation (absent, present [hypermature villi for gestational period, increase in syncytial knots (subjective and “normalized” for gestational age per the pathologist’s experience, patchy fibrin adherent to the syncytial layer, some areas with lower villous density])
2. Villitis of unknown etiology (absent, low grade [presence of inflammation in ≤10 adjacent villi in any one focus], high grade [presence of inflammation in multiple foci across sections, with inflammation in >10 adjacent villi in at least one section])
3. Branching maturity:
  a. the estimated proportion of villi that were stem villi (%), immature intermediate villi (%), mature intermediate villi (%) or terminal villi (%);
4. Stromal maturity:
  a. the estimated proportion of intervillous space that was composed of stellate reticulum (the immature myxoid stroma that developmentally preceded collagenisation) (%)
  b. the level of collagen in stem villi (none, some, a lot);
  c. the amount of smooth muscle surrounding stem vessels (none, some, a lot);
  d. Hofbauer cells (stellate reticulum is richer in Hofbauer cells than collagenized stroma; few, many);
5. Capillary maturity:
  a. vasculosyncytial membranes (none, few, many);
  b. villi capillarization (none, few, many);
6. Syncytiotrophoblast layer maturity:
  a. syncytial knots (none, few, many)
  b. syncytial thickness (thin, thick);
7. Cytotrophoblast layer maturity:
  a. cytotrophoblast density (none, thin, thick);
8. A global assessment of placental maturity relative to GA at delivery (appropriate, immature, or mature for GA).

Summary scores were calculated for three types of placental maturity (overall branching maturity, villous stromal maturity, and syncytiotrophoblast maturity; Supplementary Methods):

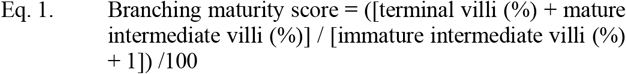

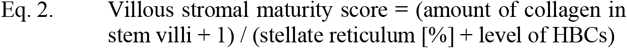

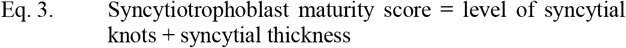

### Transcriptome profiling

To determine whether relationships exist between placental expression of gene pathways associated with NTDs and placental histopathology in cases, we evaluated relationships between placental histopathology and transcriptome signatures. Methods for placental transcriptome sequencing have been previously described and the results reported (17), and here, these data were analysed in relation to placental histopathology.

We assessed relationships between placental histopathology and: 1. overall (global) placental gene expression patterns, 2. dysregulated gene expression in cases, and 3. dysregulated expression in 14 previously identified geneset clusters of interest, that were up- or down-regulated in cases (Supplementary Methods) (17).

### Statistical analysis

Data were analysed using JMP (version 16.0, SAS Institute Inc, NC). Associations between study group and placental anthropometry were tested using unadjusted (one-way Analysis of Variance [ANOVA], for parametric data); Wilcoxon Rank Sum test, for non-parametric data) and adjusted (standard least squares regression and Tukey’s HSD post-hoc test) models. Differences in placental histopathology between study groups were assessed using unadjusted and adjusted nominal logistic regression models. All adjusted models included infant sex (male, female; categorical) and GA at delivery (20-41 weeks; continuous). Data are median (IQR) with p values from adjusted standard least squares regression, or n (%) with adjusted odds ratio (aOR [95% confidence interval (CI)]) with p values from Likelihood Ratio Chi Square test. Statistical significance was set at p<0.05.

To determine whether key maternal and infant characteristics may relate to placental histopathological phenotype, we also explored associations between: 1. GA at delivery with placental anthropometry and histopathology (generalized linear models with normal or binomial distribution; data are parameter estimate [β; 95% CI] with p value from Likelihood Ratio Chi Square test]) and 2. maternal and infant data with placental histopathology (Spearman’s rank correlation test [rho; ρ] with raw and FDR-adjusted q values).

### Assessment of placental transcriptome features that discriminate placentae based on maturity

Principal components analysis (PCA) and linear and logistic regression models were used to investigate relationships between placental transcriptome signatures and overall placental maturity (maturity scores and overall rating of maturity relative to GA), following methods applied similarly in previous studies (27-29). To determine how samples clustered based on placental transcriptome features of interest, multiple PCAs were performed as part of a data-reduction strategy, which included: 1. expression levels of the top 20% of expressed and annotated transcripts, 2. expression levels of the differentially expressed genes (DEGs) in cases compared to both PT and SB controls, and 3. expression levels of the genes associated with each of the 14 geneset clusters of interest, previously identified to be up- or downregulated in cases (Supplementary Methods). Cytoscape (version 3.8.2) was used to construct gene interaction networks (30).

## Results

### Clinical cohort characteristics

Baseline characteristics have been described elsewhere (17). In brief, maternal demographic and clinical characteristics did not differ between cases, PT controls and SB controls. *In utero* fetal spina bifida closure was performed for six cases. GA at delivery was similar between cases and PT controls, and shorter in both groups compared to SB controls (17).

### Case placentae are smaller and have higher risk of pathology and maldevelopment

Placental weight data were analysed for cases compared to PT controls (placental weights were missing for 2/12 [16.7%] cases). Compared to PT controls, cases had lower placental weight (263 g [175, 370] vs. 455 g [378, 560], p=0.001) and placental weight z-scores (−0.4 [-1.6, 0.2] vs. 1.3 [0.7, 2.2], p=0.0003), but higher birthweight-to-placental weight ratios (5.5 [3.9, 7.4] vs 4.4 [4, 4.7], p=0.002; Figure 1A, Supplementary Table S1).

**Figure 1.**
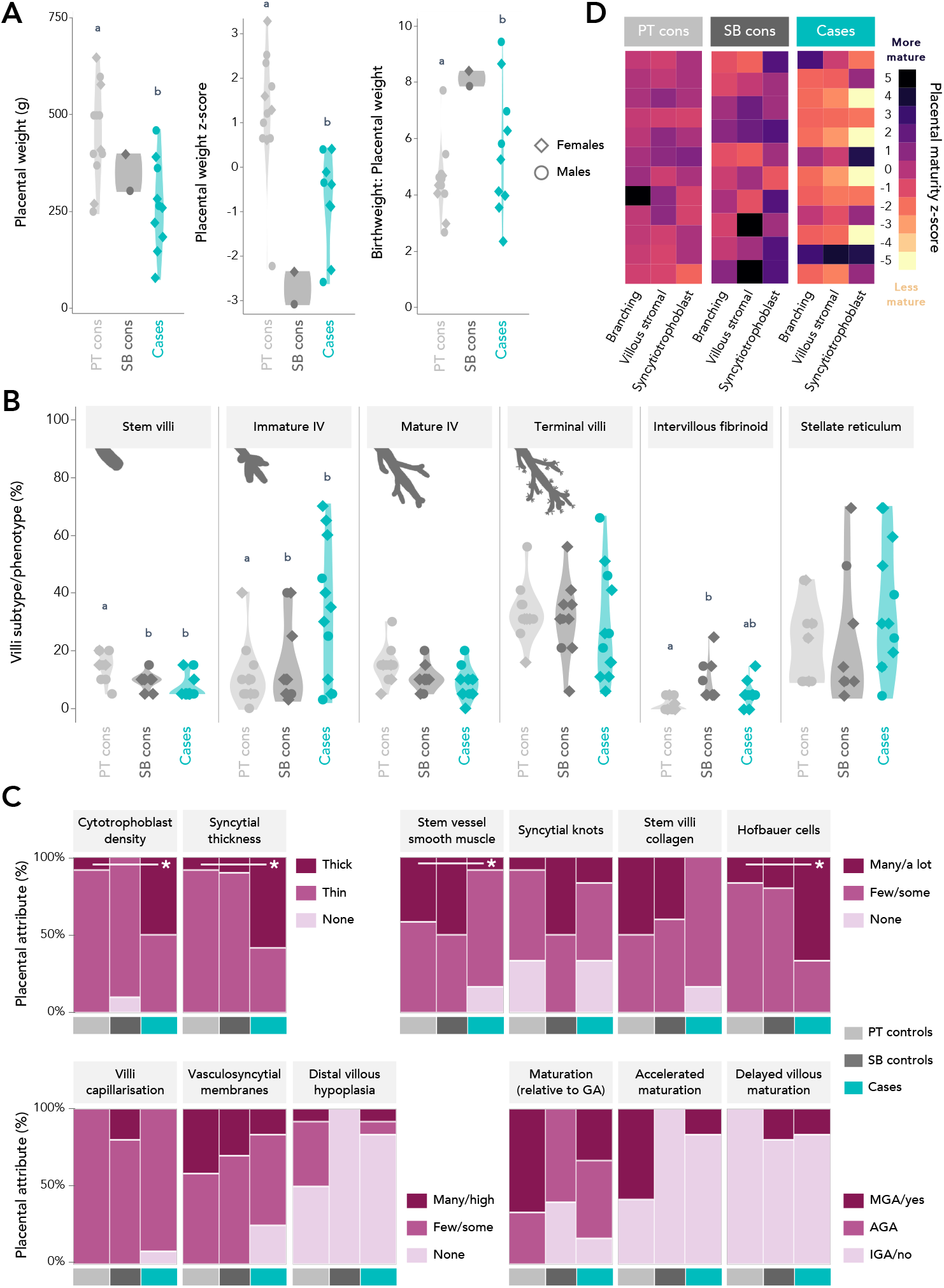
Placental anthropometry and histopathological characteristics stratified by study group. Data are plotted as violin plots (A, C; representing the density of data at different y-axis values), heat maps (B; representing standardised placental maturity scores; lighter colours=lower z-score, darker colours=higher z-score), and mosaic plots (D; proportion of participants in each group with a given placental histopathological characteristic; width of column = proportionate to the study group size). Statistical significance is denoted by different connecting letters (A, C; Tukey’s post hoc from adjusted analyses) or an asterisk (D; Likelihood Ratio Chi Square test from adjusted logistic regression). Cons = controls. IV = intermediate villi. MGA = mature for gestational age. AGA = appropriate maturity for gestational age. IGA = immature for gestational age.

Placental histopathology was assessed for all study participants. Case placentae had a higher proportion of immature intermediate villi (IIV; 32.5% [6.25, 56.3]), and a lower proportion of stem villi (5% [5, 8.8]), than PT controls (IIV: 10% [5, 13.8], p=0.01; Stem villi: 15% [11.3, 20], p=0.003; Figures 1B and S1, Supplementary Table S1). In comparison to PT controls, cases also had higher odds of having many (vs. few) HBCs (aOR=16.2 [1.44, 580], p=0.02), thick cytotrophoblast (vs. thin/no cytotrophoblast; aOR=14.9 [1.1, 609], p=0.04) and syncytiotrophoblast (vs. thin syncytiotrophoblast; aOR=146 [3, 3.5e5], p=0.007) layers, and lower odds of having a lot of smooth muscle around stem vessels (vs. some/none; aOR=0.08 [0.003, 0.9], p=0.04; Figures 1C and S1, Supplementary Table S1). Case placental histopathology did not differ from full term SB controls (Figure 1A-C, Supplementary Table S1).

In comparing measures of overall placental maturity, cases had lower syncytiotrophoblast maturity scores (3 [3, 5]) than PT (5 [4, 5]) and SB controls (5.5 [5, 6], unadjusted p=0.01), suggesting relative syncytiotrophoblast immaturity, however, the difference was not evident in adjusted analyses (Figure 1D, Supplementary Table S1).

### Global placental transcriptome signatures associate with placental histopathological maturity

In analysis of relationships between the top 10 global gene expression principal components (PCs) and measures of placental maturity (Supplementary Tables S2-7), we found that inclusive of all study groups, branching maturity scores were associated with global gene expression PC9 (β=0.001 [0.0001, 0.001], p=0.02; Table 1). PC9 was characterized by genes involved in cell cycle, DNA damage/repair and nucleic acid binding (Table 2). Syncytiotrophoblast maturity scores were associated with global gene expression PC6 (β=0.01 [0.002, 0.02], p=0.02) and PC8 (β=0.01 [0.004, 0.02], p=0.008; Table 1). PC6 was characterized by genes involved in KIT receptor signaling, which regulates placental trophoblast invasion and differentiation (31, 32), and Interleukin-5 signaling, which promotes the proliferation and differentiation of various immune cells (33) (Table 2). PC8 was characterized by genes involved in RNA metabolism and processing (Table 2).

**Table 1.**
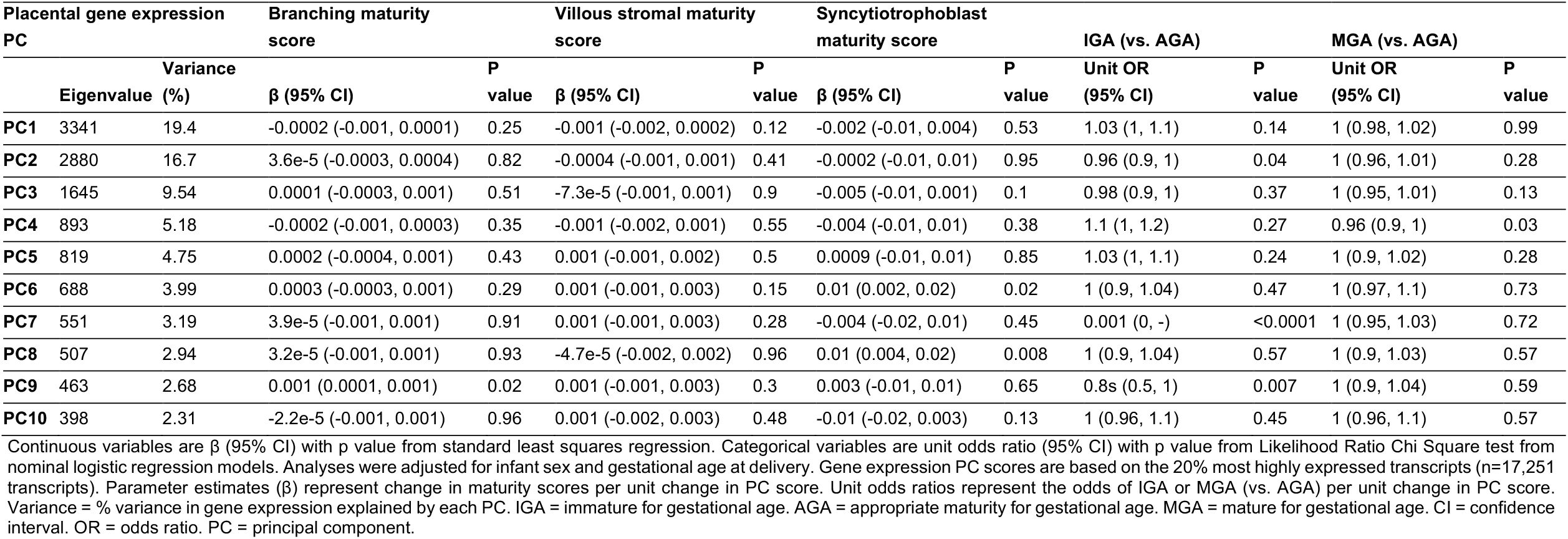
Relationships between gene expression principal component scores and placental histopathological maturity ratings.

**Table 2.**
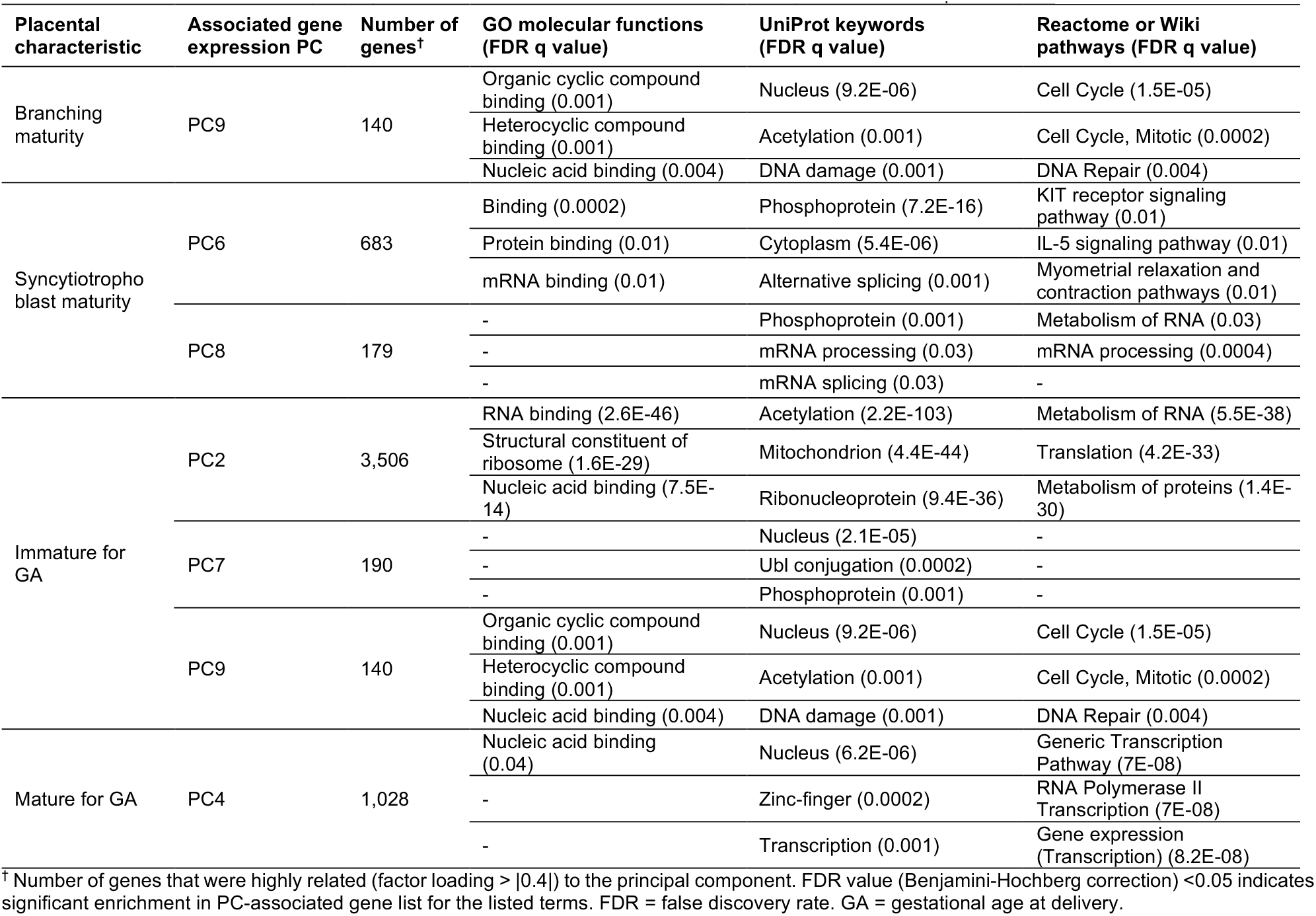
Functional enrichment analysis of gene expression principal components significantly associated with placental histopathological maturity ratings.

Placental immaturity across the cohort was associated with global gene expression PC2 (β=0.96 [0.91, 1], p=0.04), PC7 (β=0.001 [0, -], p<0.0001) and PC9 (β=0.8 [0.5, 1], p=0.007; Table 1). PC2 was characterized by genes involved in translation and RNA and protein metabolism, PC7 was enriched for genes associated with UniProt keywords “Ubl conjugation” and “phosphoprotein”, suggesting a role in post-translational protein modifications, and wile PC9 was characterized as described above by genes involved in cell cycle, DNA damage/repair and nucleic acid binding (Table 2). Conversely, overall placental hypermaturity was associated with global gene expression PC4 (β=0.96 [0.9, 1], p=0.03), which was characterized by genes involved in gene expression, and was enriched for genes associated with UniProt keyword “zinc-finger” (Table 2).

### Differentially expressed genes and gene pathways in cases associate with measures of placental histopathological maturity

In analyses of relationships between the top DEG and geneset PCs (PC1 from each PCA model) and measures of placental maturity, we found that both branching maturity scores (β=-0.02 [-0.04, -0.006], p=0.01) and villous stromal maturity scores (β=-0.03 [-0.1, - 0.01], p=0.007) in cases were associated with Hedgehog signaling PC1 (Table 3, Figure 2A-C). Villous stromal maturity scores in cases were also associated with immune and inflammatory processes PC1 (β=-0.01 [-0.02, -0.002], p=0.03; Table 3, Figure 2D-E), spinal cord injury PC1 (β=-0.02 [-0.03, -0.0004], p=0.046; Table 3, Figure 2F-G), and Wnt signaling PC1 (β=-0.03 [-0.05, -0.0005], p=0.047; Table 3, Figure 2H-I). Hedgehog and Wnt signaling, spinal cord injury and immune and inflammatory processes were each upregulated in case placentae (17). Full lists of the genes characterizing each of the 14 enriched geneset clusters in cases are provided in Supplementary Tables S8-S21.

**Table 3.** Relationships between the first principal component scores for differentially expressed genes and genesets, and placental histopathological maturity ratings in cases.

| Gene expression category | PC1 attributes |  | Branching maturity score |  | Villous stromal maturity score |  | Syncytiotrophoblast maturity score |  |
| --- | --- | --- | --- | --- | --- | --- | --- | --- |
| | Eigenvalue | Variance (%) | $\beta$ (95% CI) | P value | $\beta$ (95% CI) | P value | $\beta$ (95% CI) | P value |
| Differentially expressed genes PC1 | 268 | 68.6 | 0.002 (-0.001, 0.01) | 0.17 | 0.003 (-0.002, 0.01) | 0.23 | 0.01 (-0.04, 0.1) | 0.59 |
| Enriched geneset clusters PC1 |  |  |  |  |  |  |  |  |
| Angiogenesis | 137 | 18.5 | -0.003 (-0.01, 0.001) | 0.1 | -0.005 (-0.01, 0.001) | 0.09 | -0.03 (-0.1, 0.5) | 0.42 |
| COPII | 87 | 23.5 | 0.002 (-0.004, 0.01) | 0.47 | -0.001 (-0.01, 0.01) | 0.92 | -0.01 (-0.11, 0.1) | 0.79 |
| ECM activities | 84 | 24.4 | -0.004 (-0.01, 0.002) | 0.15 | -0.01 (-0.02, 0.002) | 0.11 | -0.04 (-0.13, 0.1) | 0.43 |
| ER activities | 37.1 | 27.7 | 0.002 (-0.01, 0.01) | 0.59 | -0.002 (-0.02, 0.01) | 0.77 | -0.02 (-0.17, 0.14) | 0.82 |
| Glycosylation | 68.8 | 24.2 | 0.002 (-0.004, 0.01) | 0.43 | -0.0001 (-0.01, 0.01) | 0.98 | -0.02 (-0.12, 0.1) | 0.72 |
| Hedgehog signaling | 13.5 | 28.6 | -0.02 (-0.04, -0.01) | 0.01 | -0.03 (-0.1, -0.01) | 0.007 | -0.2 (-0.05, 0.13) | 0.21 |
| Immune/inflammatory processes | 29.8 | 22.1 | -0.004 (-0.01, 0.004) | 0.27 | -0.01 (-0.02, -0.002) | 0.03 | -0.1 (-0.2, 0.04) | 0.16 |
| Lipid metabolism | 39.6 | 20.2 | 0.004 (-0.003, 0.01) | 0.27 | 0.002 (-0.01, 0.01) | 0.64 | -0.004 (-0.12, 0.11) | 0.95 |
| Protein synthesis | 59.7 | 31.4 | -0.002 (-0.01, 0.002) | 0.26 | -0.004 (-0.01, 0.001) | 0.13 | -0.02 (-0.1, 0.1) | 0.59 |
| SHP2 activity | 12.5 | 22 | -0.01 (-0.02, 0.01) | 0.26 | -0.01 (-0.03, 0.01) | 0.3 | 0.03 (-0.2, 0.3) | 0.74 |
| SLC transport | 12.9 | 22.7 | 0.01 (-0.01, 0.02) | 0.24 | 0.01 (-0.01, 0.03) | 0.29 | 0.1 (-0.11, 0.3) | 0.32 |
| Spinal cord injury | 18.4 | 20.2 | -0.01 (-0.02, 0.01) | 0.25 | -0.02 (-0.03, -0.0004) | 0.046 | -0.1 (-0.3, 0.1) | 0.46 |
| Transmembrane transport | 38.8 | 23.1 | -0.001 (-0.01, 0.01) | 0.87 | -0.01 (-0.02, 0.01) | 0.27 | -0.03 (-0.2, 0.1) | 0.57 |
| Wnt signaling | 14.1 | 27.6 | -0.01 (-0.03, 0.01) | 0.14 | -0.03 (-0.1, -0.001) | 0.047 | -0.2 (-0.5, 0.1) | 0.13 |

Data are $\beta$ (95% CI) with p value from standard least squares regression. Analyses were adjusted for infant sex and gestational age at delivery. Parameter estimates ( $\beta$ ) represent change in maturity scores per unit change in PC score. Variance = % variance in gene expression explained by the first PC. CI = confidence interval. PC = principal component. COPII = Coat Protein Complex II. ECM = extracellular matrix. SHP2 = Src-homology 2 domain-containing phosphatase 2. SLC = solute carrier.

**Figure 2.**
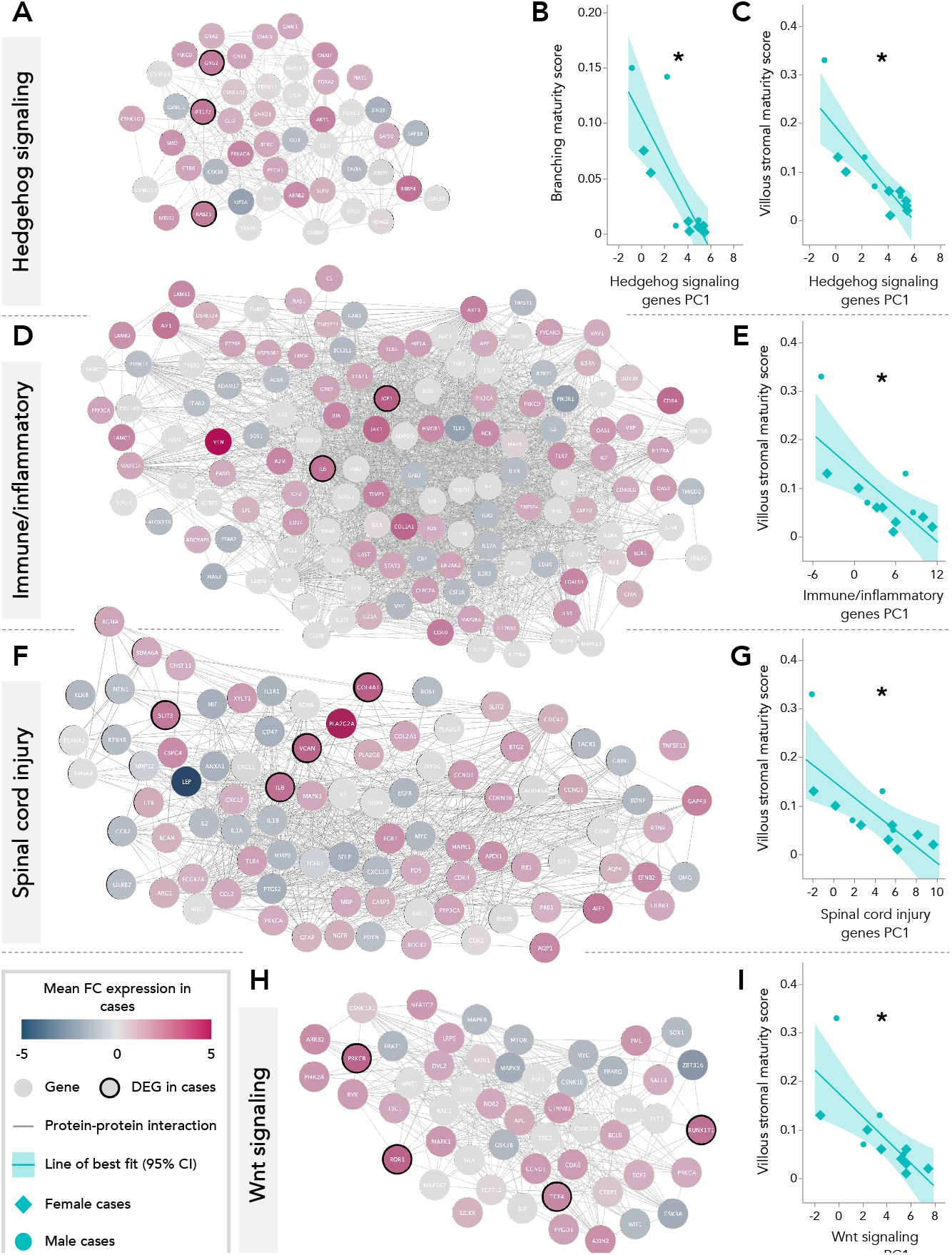
Gene-interaction networks for genes characterizing enriched geneset clusters associated with placental histopathological maturity in cases. A. Expression patterns in cases of genes involved in Hedgehog signaling, which was upregulated in case placentae. B-C. Hedgehog signaling PC1 was associated with branching (β=-0.02 [-0.04, -0.006], p=0.01) and villous stromal (β=-0.03 [-0.1, -0.01], p=0.007) maturity scores in cases. D. Expression patterns in cases of genes involved in immune and inflammatory processes, which were upregulated in case placentae. E. Immune and inflammatory processes PC1 was associated with villous stromal maturity scores in cases (β=-0.01 [-0.02, -0.002], p=0.03). F. Expression patterns in cases of genes involved in the spinal cord injury response pathway, which was upregulated in case placentae. G. Spinal cord injury PC1 was associated with villous stromal maturity scores in cases (β=-0.02 [-0.03, -0.0004], p=0.046). H. Expression patterns in cases of genes involved in Wnt signaling, which was upregulated in case placentae. I. Wnt signaling PC1 was associated with villous stromal maturity scores in cases (β=-0.03 [-0.05, -0.0005], p=0.047). Circles in network diagrams = genes, edges = connection between interacting genes (proteins). PC = principal component. FC = fold change. DEG = differentially expressed gene. Full lists of the genes characterizing each of the 14 enriched geneset clusters in cases are provided in Supplementary Tables S8-S21.

### Gestational age influences placental characteristics overall and within groups

In the full cohort, GA at delivery was positively associated with placental weight, birthweight-to-placental weight ratio, proportion of placental villi that were terminal villi, levels of vasculosyncytial membranes and syncytial knots, amount of collagen in stem villi and smooth muscle around stem vessels, and overall villous stromal and syncytiotrophoblast maturity scores (Table 4, Supplementary Figures S2 and S3). GA at delivery was negatively associated with the amount of stellate reticulum contained in the placental villi, proportion of placental villi that were immature intermediate villi, number of HBCs, cytotrophoblast density, and syncytiotrophoblast thickness (Table 4). Most of these associations were also observed within cases when stratified by study group, with a few exceptions: there was no association between GA at delivery and villous stromal maturity in cases, and the sample sizes for a response level of four of the pathology variables in cases were too small for comparisons (Table 4).

**Table 4.**
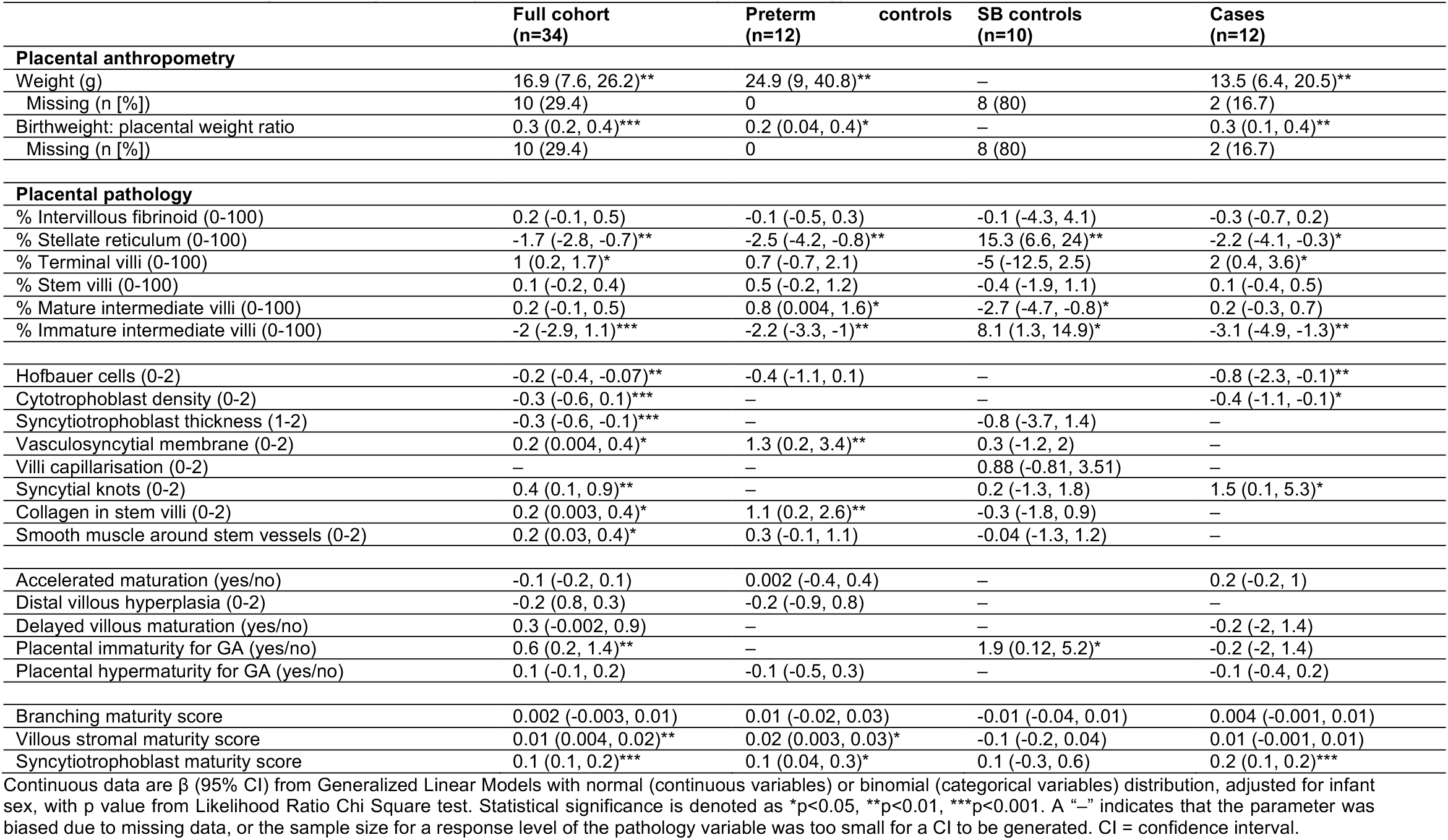
Associations between gestational age at delivery and placental anthropometry and pathology in cases and controls.

Case placentae demonstrated attributes associated with a younger GA (higher cytotrophoblast density and syncytial thickness, higher levels of HBCs, larger proportion of placental villi composed of stellate reticulum, and larger proportion immature intermediate villi; Figure 3). Concurrently, case placentae also had attributes associated with an older GA and villous hypermaturity (similar proportion of mature intermediate villi, vasculosyncytial membranes, and syncytial knots as full-term born SB controls; Figure 3).

**Figure 3.**
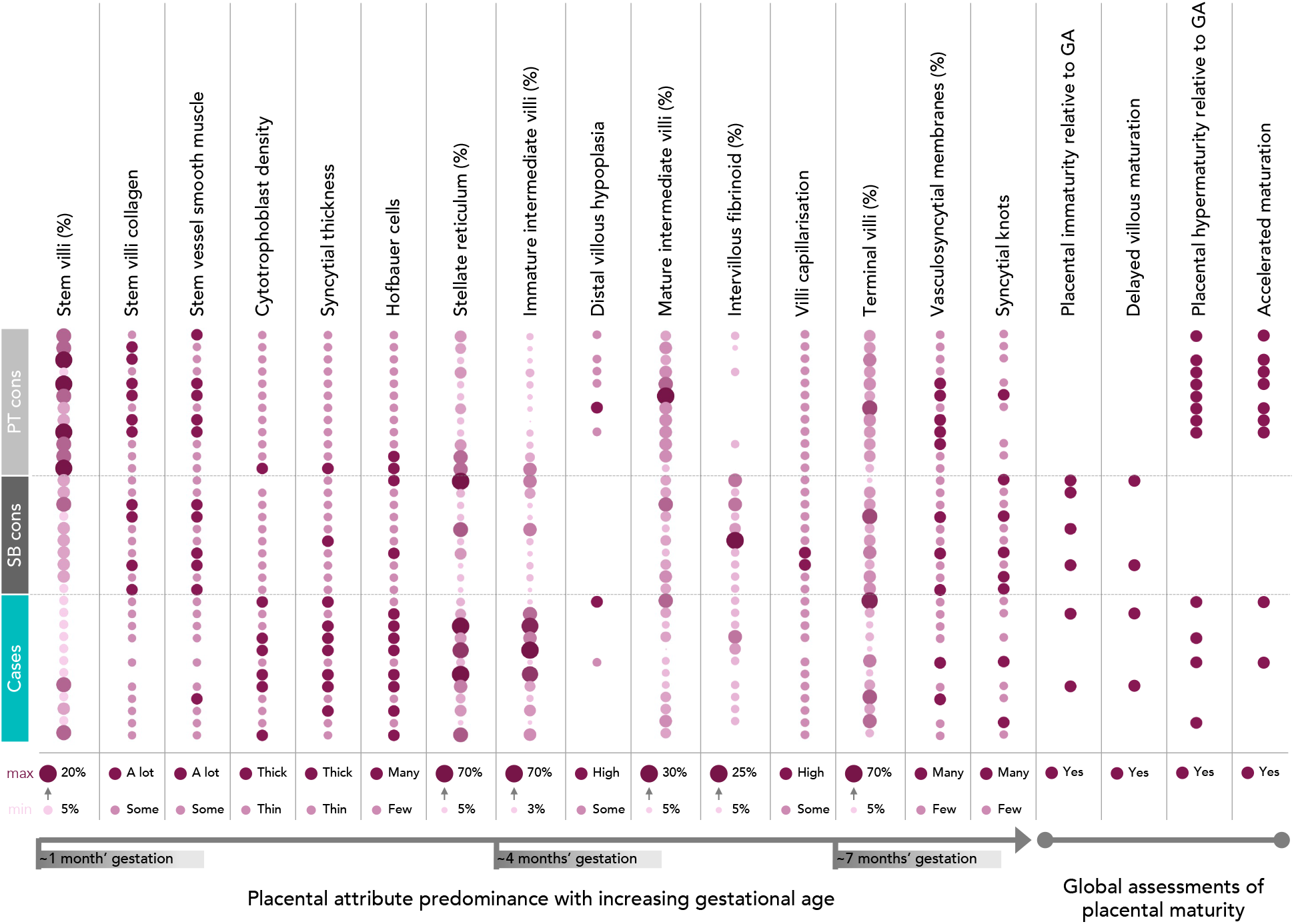
Placental histopathological characteristics ordered (left to right) by approximate emergence across gestational age. Larger circles/darker colour = more/higher proportion of that attribute. Smaller circles/lighter colour = less/lower proportion of that attribute. No circle = not observed. A lot/some, Thick/thin, Many/few = categorical outcome variables. Numbers (%) with arrows = continuous outcome variables.

### Case placental histopathology differs by fetal sex

Male cases had a lower proportion of intervillous fibrinoid (5% [5, 5]) than male SB controls (12.5% [10, 15]), but a higher proportion than male PT controls (0% [0, 3.8], p=0.0005; Supplementary Table S22). The proportion of placental villi that were stem villi was also lower in male cases than male PT controls (5 [5, 12.5] vs. 15 [10, 18.8], p=0.04; Supplementary Table S22).

Female cases had lower placental weight (221 g [147, 283] vs. 495 g [305, 633], p=0.01) and placental weight z-scores (−0.6 [-1.2, 0.03] vs. 1.5 [0.8, 2.9], p=0.007) than female PT controls (Supplementary Table S23). Birthweight-to-placental weight ratio was also associated with study group among female fetuses, however, *post-hoc* analyses revealed no group differences (Supplementary Table S23). The proportion of placental villi that were stem villi was also lower in female cases than female PT controls (5 [5, 8.8] vs. 17.5 [15, 20], p=0.0002; Supplementary Table S23).

### Maternal clinical and dietary characteristics associate with placental histopathology

There were no associations between placental maturity scores and summary measures of maternal dietary intake in the full cohort, or in cases or SB controls after stratification by study group (Supplementary Figure S4). Although there were some associations between individual dietary components and placental histopathology in the full cohort and among cases and SB controls separately, after FDR correction, only an association between maternal reported folate intake and the proportion of villi that were mature intermediate villi in SB controls remained (FDR p value=0.02; Figure 4).

**Figure 4.**
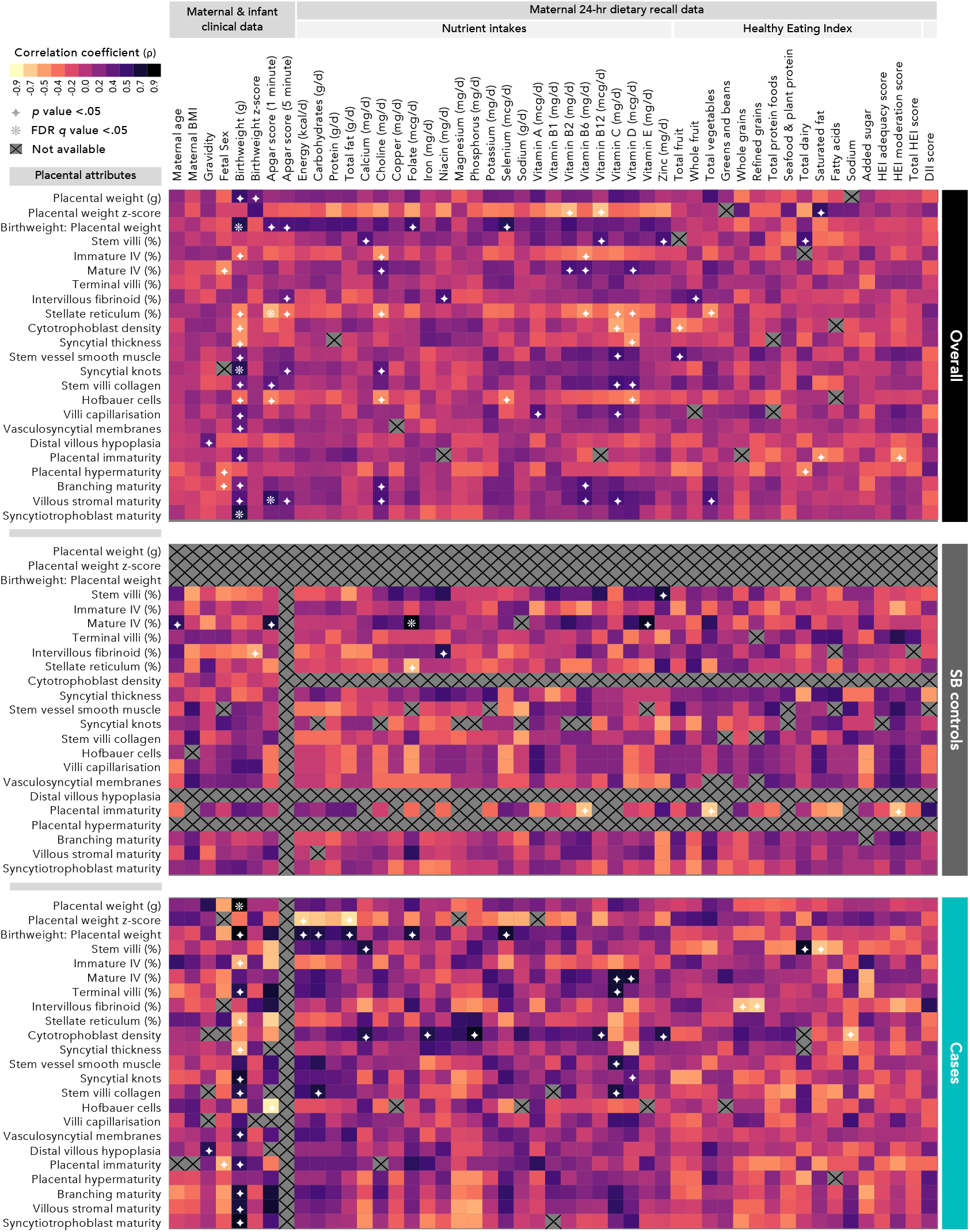
Associations between maternal and infant clinical data, maternal dietary recall data, and placental histopathological characteristics. Data are from Spearman’s rank correlation (rho; ρ) test with raw and false discovery rate (FDR)-adjusted q values. Spearman’s ρ are represented in heatmaps (yellow-to-orange colours=negative correlation, pink-to-black colours=positive correlation). Statistical significance is denoted by white symbols (✦ = raw p value <0.05; ❋ = FDR q value <0.05). Grey squares with X = data unavailable.

## Discussion

We have shown for the first time in a contemporary cohort that isolated fetal open SB associates with low placental weight and an increased risk of placental pathology, in comparison to a control group matched to cases for GA and maternal pre-pregnancy BMI. We also found that dysregulated placental expression of gene pathways implicated in NTDs associates with placental histopathology in cases, suggesting that placental maldevelopment in these pregnancies may be shaped by a uterine environment that is also permissive of NTDs. These findings improve our understanding of placental maldevelopment in pregnancies with fetal NTDs, and the mechanisms that may underly NTD-associated comorbidities, such as poor fetal growth and development and early birth.

Despite that all but two cases were born preterm, case placentae had similar levels of mature intermediate and terminal villi, vasculosyncytial membranes, and syncytial knots, as the full term-born SB controls. Premature villous differentiation towards terminal villi can diminish the placenta’s capacity to grow (34, 35), which may have led to the low placental weight observed in cases. Villous hypermaturity and increased syncytial knots in preterm placentae are associated with uteroplacental malperfusion (36, 37), and may result from oxidative damage (38, 39). Notably, expression levels of catalase (*CAT*), a key antioxidant enzyme that responds to oxidative stress (40), were increased in case placentae, and *CAT* was predicted to regulate 20% of the differentially expressed genes in cases (17). Case placentae also had higher HBC levels than controls, which are known to express *CAT* in early gestation and have a role in oxidative stress generation and regulation (41, 42). Decreased maternal antioxidant capacity has also been reported in pregnancies with fetal NTDs (43). Taken together, it is possible that upregulated placental *CAT* expression is an adaptive response to increased oxidative stress levels in cases, for which villous hypermaturity and increased syncytial knots may be histologic markers.

Case placentae also displayed characteristics of functional and cellular immaturity, including a higher proportion of immature intermediate villi and HBC levels, and a thicker syncytial layer, than the GA-matched PT controls. The elevated HBC levels in cases are consistent with findings from our previous work, where placentae from fetuses with isolated NTDs had higher HBC levels and an increased risk of villous hypermaturity, in comparison to a well-matched control group (4). HBCs are folate-dependent fetal macrophages that regulate placental vasculogenesis, angiogenesis, and inflammation (44, 45). We previously posited that suboptimal maternal folate status in case pregnancies may have led to HBC dysregulation, and subsequent placental pathologies, given that the period of CPP recruitment (1959-1965) preceded folic acid fortification and supplementation (4). However, as folate deficiency in the current day Canadian population is rare (16), it is plausible that the increase in HBC levels in case placentae is due to other reasons, such as an adaptation following low bioavailability of nutrients required for folate metabolism (vitamins B6, B12, and choline (46, 47)), or maternal immune dysregulation (48). In agreement, mothers of cases in this cohort reported lower intakes of vitamins B6 and B12 and choline than controls (17). Whether isolated fetal SB may associate with a shift from anti-to pro-inflammatory HBC phenotypes (48), and how this may influence placental development and function, is an area requiring further investigation.

We also found that in cases, placental villous stromal maturity was associated with expression patterns in upregulated immune and inflammatory process gene pathways (17). Our summary measure of villous stromal maturity considered HBC levels and the amount of collagen and stellate reticulum. Immune dysregulation and increased inflammatory load in pregnancy are associated with an increased risk of fetal SB (49, 50), and upregulation of these inflammation-related pathways may have induced HBC proliferation, as has been observed in other pro-inflammatory pregnancy states (51). Placental villous stromal maturity in cases was also associated with gene expression in the spinal cord injury response pathway (17), characterized by pro-inflammatory secondary lesion cascades that are activated following improper neural tube closure (52). Together, these findings suggest that upregulation in inflammation-related gene processes in case placentae may associate with placental stromal cell composition, which are critical regulators of placental villous development (53). Clarifying the factors that lead to upregulation of placental inflammatory signaling in cases and resulting histologic phenotypes is necessary to reduce the risk of fetal morbidity and mortality known to associate with placental inflammation (54).

Gene expression patterns in two central regulators of development, the Hedgehog and Wnt signaling pathways (55, 56), were also associated with placental villous stromal and branching maturity scores in cases. Signaling in Hedgehog and Wnt pathways coordinates proper neural tube patterning and closure (55, 56), and disruptions in these pathways have been associated with NTDs (55, 56). In the placenta, the Hedgehog pathway modulates trophoblast syncytialization (57), while the Wnt pathway regulates trophoblast invasion and differentiation (58). Our finding that both signaling pathways were upregulated in case placentae suggests that known NTD-associated genetic signatures may be detectable in placentae from affected pregnancies (17). And here, we have shown for the first time that altered activity in these pathways may be associated with placental histologic phenotype in cases. As dysregulation in gene pathways previously linked to NTDs may associate with histologic features in the placenta, it is also possible that further study of placental histopathology in cases could provide opportunities to identify novel mechanisms associated with NTD phenotype or pathophysiology.

When stratified by fetal sex, we found that female, but not male, cases had lower placental weight than PT controls. This may suggest that there are female-specific adaptations to the uterine environment in SB-affected pregnancies, consistent with sex differences in placental adaptations to other suboptimal pregnancy exposures (59, 60). However, these differences may have been detected only in females, and not males, because the sample size for female cases was larger (as expected, given NTDs are more prevalent in females (61)), limiting our ability to draw conclusions. We also did not find any associations between maternal dietary intakes and placental histopathology in cases after correcting for FDR. Nevertheless, given the known influence of maternal nutrition on fetal NTD risk (7) and placental (mal)development and function (62), further investigation into relationships between maternal nutritional status, in larger cohorts or using circulating nutrient metabolomic assessments, and placental phenotype in NTD-affected pregnancies is warranted.

This study is the first to characterize placental histopathology, with integration of transcriptomic signatures to evaluate relationships between placental gene expression and histopathology, in a contemporary cohort of fetuses with isolated SB. We report that placentae from fetuses with isolated SB may present with histologic markers of oxidative stress and increased inflammation, and that these pathologies may be driven by dysregulation in antioxidant- and immune/inflammation-related gene networks (17). We also validate findings of elevated HBC levels and placental villous hypermaturity in cases from our previous work using the CPP data, suggesting some consistency in case placental phenotype across populations and time periods (4). An improved understanding of histopathological and genetic phenotypes in case placentae increases our understanding of the mechanistic factors that may contribute to fetal comorbidities in case pregnancies, and is ultimately needed to reduce offspring morbidity and mortality.

## Data Availability

All data produced in the present study are available upon reasonable request to the authors.

## Acknowledgements

We would like to thank Vagisha Pruthi for overseeing the study recruitment, data entry and sample collection, and the staff at the Research Centre for Women’s and Infants’ Health (RCWIH) BioBank for collecting and processing the placental samples,

This research is supported by the Canadian Institutes of Health Research (CIHR PJT-175161) and departmental research grants from the Department of Obstetrics and Gynaecology at Mount Sinai Hospital, Toronto, and the Department of Health Sciences at Carleton University, Ottawa. MW was supported by an Ontario Graduate Scholarship, Faculty of Science, Carleton University.

## Author contributions

MW, KLC, DG, and TVM conceptualized and designed the study. TVM oversaw patient recruitment and sample collection. MW, DG, JAP, and KLC carried out data collection, analysis, and interpretations. MW generated the figures. MW and KLC wrote the original manuscript draft, and all authors were involved in editing the paper and approved the final version. KLC provided supervision and KLC and TVM provided funding.

