## Supplementary Tables for "Altered placental phenotype and increased risk of placental pathology in fetal spina bifida: a matched case-control study"

**Supplementary Table S1.** Associations between isolated spina bifida and placental characteristics and pathology in cases and controls.

| | PT controls (n=12) | SB controls (n=10) | Cases (n=12) | Unadjusted p value | Adjusted p value [†] |
|---|---|---|---|---|---|
| **Placental anthropometry** | | | | | |
| Weight (g) | 455 (378, 560) [a] | 352 (304, 399) [b] | 263 (175, 370) [b] | 0.006 | 0.001 |
| Missing (n [%]) | 0 | 8 (80) | 2 (16.7) | | |
| zWeight [*] | 1.3 (0.7, 2.2) [a] | -2.7 (-3.1, -2.4) [b] | -0.4 (-1.6, 0.2) [b] | 0.0003 | 0.0003 |
| Missing (n [%]) | 0 | 8 (80) | 3 (25) | | |
| Birthweight: placental weight ratio | 4.4 (4, 4.7) [a] | 8.1 (7.8, 8.4) [ab] | 5.5 (3.9, 7.4) [b] | 0.1 | 0.002 |
| Missing (n [%]) | 0 | 8 (80) | 2 (16.7) | | |
| **Placental histopathology** | | | | | |
| % Intervillous fibrinoid | 0 (0, 4.3) [a] | 5 (5, 15) [b] | 5 (5, 5) [ab] | 0.0004 | 0.003 |
| % Stellate reticulum | 25 (10, 30) | 10 (8.8, 35) | 30 (16.3, 57.5) | 0.13 | 0.45 |
| % Terminal villi | 30 (30, 35) | 30 (20, 36.3) | 22.5 (11.3, 43.8) | 0.7 | 0.32 |
| % Stem villi | 15 (11.3, 20) [a] | 10 (8.8, 10) [b] | 5 (5, 8.8) [b] | 0.0008 | 0.0003 |
| % Mature intermediate villi | 15 (11.3, 15) | 10 (8.8, 11.3) | 10 (5, 10) | 0.02 | 0.09 |
| % Immature intermediate villi | 10 (5, 13.8) [a] | 10 (5, 28.8) [b] | 32.5 (6.3, 56.3) [b] | 0.1 | 0.01 |
| Branching maturity score | 0.05 (0.03, 0.1) | 0.04 (0.01, 0.1) | 0.01 (0.003, 0.1) | 0.18 | 0.58 |
| Villous stromal maturity score | 0.1 (0.06, 0.3) | 0.18 (0.1, 0.3) | 0.06 (0.03, 0.1) | 0.11 | 0.66 |
| Syncytiotrophoblast maturity score | 3 (2, 3) | 3.5 (3, 4) | 2 (1, 3) | 0.03 | 0.75 |

| | | | | Odds of placental pathology in cases (n=12) | | | |
|---|---|---|---|---|---|---|---|
| | PT controls (n=12) | SB controls (n=10) | Cases (n=12) | (vs. PT cons) | | (vs. SB cons) | |
| **Placental pathology** | | | | OR (95% CI) | aOR (95% CI)[†] | OR (95% CI) | aOR (95% CI)[†] |
| Hofbauer cells (n [%]) | | | | 10 (1.7, 89.8)* | 16.2 (1.4, 580)* | 8 (1.3, 73.1)* | 0.75 (0.03, 14.9) |
| Many | 2 (16.7) | 2 (20) | 8 (66.7) | | | | |
| Few | 10 (83.3) | 8 (80) | 4 (33.3) | | | | |
| Cytotrophoblast density (n [%]) | | | | 11 (1.42, 235)* | 14.9 (1.12, 609)* | - | - |
| Thick | 1 (8.33) | 0 | 6 (50) | | | | |
| Thin/None | 11 (91.7) | 10 (100) | 6 (50) | | | | |
| Syncytiotrophoblast density (n [%]) | | | | 15.4 (2, 332)** | 146 (2.95, 3.46e5)** | 12.6 (1.6, 274)* | 0.14 (0.0005, 11) |
| Thick | 1 (8.3) | 1 (10) | 7 (58.3) | | | | |
| Thin | 11 (91.7) | 9 (90) | 5 (41.7) | | | | |
| Vasculosyncytial membrane (n [%]) | | | | 0.3 (0.03, 1.7) | 0.09 (0.003, 1.2) | 0.47 (0.05, 3.5) | 22.5 (0.8, 1304) |
| Many | 5 (41.7) | 3 (30) | 2 (16.7) | | | | |
| Few/None | 7 (58.3) | 7 (70) | 10 (83.3) | | | | |
| Villi capillarisation (n [%]) | | | | - | - | - | - |
| Many | 0 | 2 (20) | 0 | | | | |
| Few/None | 12 (100) | 8 (80) | 12 (100) | | | | |
| Syncytial knots (n [%]) | | | | 2.2 (0.18, 51.7) | 1.69 (0.1, 51.1) | 0.2 (0.02, 1.3) | 6 (0.2, 369) |
| Many | 1 (8.3) | 5 (50) | 2 (16.7) | | | | |
| Few/None | 11 (91.7) | 5 (50) | 10 (83.3) | | | | |

| | | | | | | | |
|---|---|---|---|---|---|---|---|
| Collagen in stem villi (n [%]) | | | | - | - | - | - |
| Lots | 6 (50) | 4 (40) | 0 | | | | |
| Some/None | 6 (50) | 6 (60) | 12 (100) | | | | |
| Smooth muscle around stem vessels (n [%]) | | | | 0.13 (0.006, 1) | 0.1 (0.003, 0.9)* | 0.1 (0.004, 0.8)* | 0.8 (0.02, 18) |
| Lots | 5 (41.7) | 5 (50) | 1 (8.3) | | | | |
| Some/None | 7 (58.3) | 5 (50) | 11 (91.7) | | | | |
| Accelerated maturation (n [%]) | | | | 0.1 (0.02, 0.9)* | 0.2 (0.02, 1.1) | - | - |
| Yes | 7 (58.3) | 0 | 2 (16.7) | | | | |
| No | 5 (41.7) | 10 (100) | 10 (83.3) | | | | |
| Distal villous hypoplasia (n [%]) | | | | 0.2 (0.02, 1.2) | 0.2 (0.02, 1.5) | - | - |
| Any | 6 (50) | 0 | 2 (16.7) | | | | |
| None | 6 (50) | 10 (100) | 10 (83.3) | | | | |
| Delayed villous maturation (n [%]) | | | | - | - | 0.8 (0.1, 7) | 7 (0.11, 457) |
| Yes | 0 | 2 (20) | 2 (16.7) | | | | |
| No | 12 (100) | 8 (80) | 10 (83.3) | | | | |
| Maturation (n [%]) ‡ | | | | | | | |
| Immature | 0 | 4 (40) | 2 (16.7) | - | - | 0.5 (0.05, 3.7) | 89.3 (0.3, 9.6e5) |
| Appropriate | 4 (33.3) | 6 (60) | 6 (50) | | | | |
| Hypermature | 8 (66.7) | 0 | 4 (33.3) | 0.3 (0.1, 1.8) | 0.9 (0.1, 9.5) | - | - |

† Adjusted for infant sex and gestational age at delivery.

‡ Reference group: appropriate maturity.

Continuous data are median (IQR) with p value for one-way analysis of variance (Unadjusted: ANOVA [normal distribution/equal variance] or Wilcoxon test [non-parametric data or outliers]; Adjusted: Multiple linear regression models). Categorical data are n (%) and odds ratios or adjusted odds ratios (95% CI) for presence of pathology in case placentae (reference groups: PT controls and SB controls) from Nominal Logistic regression models with p value from Likelihood Ratio Chi Square test. Statistical significance is denoted as *p<.05, **p<.01, ***p<.001. CI = confidence interval. OR = odds ratio. aOR = adjusted odds ratio.

**Supplementary Table S22.** Associations between isolated spina bifida and infant birth outcomes and placental phenotype in male cases and controls.

| **Infant outcomes at delivery** | **Preterm controls (n=8)** | **SB controls (n=2)** | **Cases (n=4)** | **Unadjusted p value** | **Adjusted p value [†]** |
|---|---|---|---|---|---|
| Stillborn/TOP (n [%]) | - | - | 2 (50) | - | - |
| GA at birth (weeks) [‡] | 33.2 (31.3, 34.6) | 39 (37.9, 40) | 34.8 (33.5, 36.5) | 0.03 | - |
| <37 weeks GA at birth (n [%]) | 8 (100) | 0 | 3 (75) | 0.007 | - |
| Sub-categories of PT birth (n [%]) | | | | 0.06 | - |
| Moderate/late PT (32-36 weeks GA) | 5 (62.5) | 0 | 3 (75) | | |
| Very PT (28-32 weeks GA) | 2 (25) | 0 | 0 | | |
| Extremely PT (<28 weeks GA) | 1 (12.5) | 0 | 0 | | |
| Birthweight z score | 0.32 (-0.35, 0.46) | -1.81 (-2.07, -1.54) | -0.1 (-0.24, 1.03) | 0.007 | - |
| Apgar at one minute (/10) | 9 (4, 9) | 6.5 (4, 9) | 8 (7, 9) | 0.94 | 0.24 |
| Apgar score <7 (n [%]) | 3 (37.5) | 1 (50) | 0 | 0.23 | 0.08 |
| Missing (n [%]) | 1 (12.5) | 0 | 1 (25) | | |
| Apgar at five minutes (/10) | 9 (8, 9) | 9 (9, 9) | 9 (9, 9) | 0.27 | 0.41 |
| Apgar score <7 (n [%]) | 0 | 0 | 0 | - | - |
| Missing (n [%]) | 1 (12.5) | 0 | 1 (25) | | |
| **Placental anthropometry** | | | | | |
| Weight (g) | 450 (378, 500) | 304 | 363 (265, 461) | 0.37 | 0.24 |
| Missing (n [%]) | 0 | 1 (50) | 1 (25) | | |
| zWeight [*] | 1.12 (0.66, 2.23) | -3.08 | -0.34 (-2.58, 0.41) | 0.047 | - |
| Missing (n [%]) | 0 | 1 (50) | 1 (25) | | |
| Birthweight: placental weight ratio | 4.58 (4.1, 5.25) | 7.83 | 6.94 (5.79, 9.4) | 0.06 | 0.18 |
| Missing (n [%]) | 0 | 1 (50) | 1 (25) | | |
| **Placental pathology** | | | | | |
| % Intervillous fibrinoid | 0 (0, 3.75) [a] | 12.5 (10, 15) [b] | 5 (5, 5) [c] | 0.01 | 0.0005 |
| % Stellate reticulum | 17.5 (10, 30) | 30 (10, 50) | 20 (7.5, 36.3) | 0.85 | 0.55 |
| % Terminal villi | 32.5 (30, 38.8) | 25 (20, 30) | 35 (21.3, 60) | 0.5 | 0.66 |
| % Stem villi | 15 (10, 18.8) [a] | 12.5 (10, 15) [ab] | 5 (5, 12.5) [b] | 0.18 | 0.04 |
| % Mature intermediate villi | 15 (15, 18.8) | 12.5 (5, 20) | 12.5 (6.25, 18.8) | 0.63 | 0.58 |
| % Immature intermediate villi | 7.5 (5, 13.8) | 22.5 (5, 40) | 15 (3.5, 40) | 0.71 | 0.81 |
| Branching maturity score | 0.07 (0.03, 0.11) | 0.04 (0.006, 0.08) | 0.08 (0.008, 0.15) | 0.58 | 0.82 |
| Villous stromal maturity score | 0.18 (0.06, 0.27) | 0.16 (0.04, 0.27) | 0.1 (0.06, 0.28) | 0.9 | 0.48 |
| Syncytiotrophoblast maturity score | 3 (2.25, 3) | 3 (3, 3) | 2.5 (2, 3.75) | 0.85 | 0.2 |

| | | | | **Odds of placental pathology in cases (n=4)** | | | |
|---|---|---|---|---|---|---|---|
| | **PT controls (n=8)** | **SB controls (n=2)** | **Cases (n=4)** | **(vs. PT cons)** | | **(vs. SB cons)** | |
| **Placental pathology** | | | | **OR (95% CI)** | **aOR (95% CI)[†]** | **OR (95% CI)** | **aOR (95% CI)[†]** |
| Hofbauer cells (n [%]) | | | | | | | |
| Many | 1 (12.5) | 0 | 2 (50) | 7 (0.45, 211) | 10.9 (0.43, 1120) | - | - |

| | | | | | | | |
|---|---|---|---|---|---|---|---|
| Few | 7 (87.5) | 2 (100) | 2 (50) | | | | |
| Cytotrophoblast density (n [%]) | | | | | | | |
| Thick | 0 | 0 | 2 (50) | - | - | - | - |
| Thin/None | 8 (100) | 2 (100) | 2 (50) | | | | |
| Syncytiotrophoblast density (n [%]) | | | | | | | |
| Thick | 0 | 0 | 2 (50) | - | - | - | - |
| Thin | 8 (100) | 2 (100) | 2 (50) | | | | |
| Vasculosyncytial membrane (n [%]) | | | | | | | |
| Many | 4 (50) | 0 | 0 | - | - | - | - |
| Few/None | 4 (50) | 2 (100) | 4 (100) | | | | |
| Villi capillarisation (n [%]) | | | | | | | |
| Many | 0 | 0 | 0 | - | - | - | - |
| Few/None | 8 (100) | 2 (100) | 4 (100) | | | | |
| Syncytial knots (n [%]) | | | | | | | |
| Many | 1 (12.5) | 0 | 1 (25) | - | - | - | - |
| Few/None | 7 (87.5) | 2 (100) | 3 (75) | | | | |
| Collagen in stem villi (n [%]) | | | | | | | |
| Lots | 4 (50) | 1 (50) | 0 | - | - | - | - |
| Some/None | 4 (50) | 1 (50) | 4 (100) | | | | |
| Smooth muscle around stem vessels (n [%]) | | | | | | | |
| Lots | 3 (37.5) | 1 (50) | 0 | - | - | - | - |
| Some/None | 5 (62.5) | 1 (50) | 4 (100) | | | | |
| Accelerated maturation (n [%]) | | | | | | | |
| Yes | 5 (62.5) | 0 | 1 (25) | 0.2 (0.008, 2.43) | 0.59 (0.02, 13.5) | - | - |
| No | 3 (37.5) | 2 (100) | 3 (75) | | | | |
| Distal villous hyperplasia (n [%]) | | | | | | | |
| Any | 4 (50) | 0 | 1 (25) | 0.33 (0.01, 4.05) | 0.93 (0.03, 23.1) | - | - |
| None | 4 (50) | 2 (100) | 3 (75) | | | | |
| Delayed villous maturation (n [%]) | | | | | | | |
| Yes | 0 | 0 | 2 (50) | - | - | - | - |
| No | 8 (100) | 2 (100) | 2 (50) | | | | |
| Maturation (n [%]) ‡ | | | | | | | |
| Immature | 0 | 1 (50) | 2 (50) | - | - | - | - |
| Appropriate | 2 (25) | 1 (50) | 0 | | | | |
| Hypermature | 6 (75) | 0 | 2 (50) | - | - | - | - |

† Adjusted for gestational age at delivery.
‡ Reference group: appropriate maturity.

Continuous data are median (IQR) with p value for one-way analysis of variance (Unadjusted: ANOVA [normal distribution/equal variance] or Wilcoxon test [non-parametric data or outliers]; Adjusted: Standard Least Squares models). Group differences from adjusted post-hoc tests (Tukey HSD) are indicated by different subscript letters. No letters = no group differences. Categorical data are n (%) and odds ratios or adjusted odds ratios (95% CI) for presence of pathology in case placentae (reference groups: PT controls and SB controls) from Nominal Logistic regression models with p value from Likelihood Ratio Chi Square test. A '-' indicates that data were unavailable. Statistical significance is denoted as *p<.05, **p<.01, ***p<.001. CI = confidence interval. PT = preterm. SB = spina bifida. TOP = termination of pregnancy. OR = odds ratio. aOR = adjusted odds ratio.

**Supplementary Table S23.** Associations between isolated spina bifida and infant birth outcomes and placental phenotype in female cases and controls.

| **Infant outcomes at delivery** | **Preterm controls (n=4)** | **SB controls (n=8)** | **Cases (n=8)** | **Unadjusted p value** | | **Adjusted p value** [†] | |
|---|---|---|---|---|---|---|---|
| Stillborn/TOP (n [%]) | - | 1 (12.5) | 1 (12.5) | - | | - | |
| GA at birth (weeks) [‡] | 33.7 (26.8, 35.9) | 39.7 (38.9, 40.2) | 27.2 (22.5, 35) | 0.0009 | | - | |
| <37 weeks GA at birth (n [%]) | 4 (100) | 0 | 7 (87.5) | 0.0001 | | - | |
| Sub-categories of PT birth (n [%]) | | | | 0.0007 | | - | |
| Moderate/late PT (32-36 weeks GA) | 2 (50) | 0 | 1 (12.5) | | | | |
| Very PT (28-32 weeks GA) | 1 (25) | 0 | 2 (25) | | | | |
| Extremely PT (<28 weeks GA) | 1 (25) | 0 | 4 (50) | | | | |
| Birthweight z score | 0.1 (-0.73, 0.99) | 0.16 (-0.5, 0.45) | -0.58 (-0.71, 0.17) | 0.3 | | - | |
| Apgar at one minute (/10) | 7.5 (5.5, 8) | 9 (9, 9) | 8 (4, 9) | 0.08 | | 0.16 | |
| Apgar score <7 (n [%]) | 1 (25) | 1 (12.5) | 1 (12.5) | 0.72 | | 0.27 | |
| Missing (n [%]) | 0 | 0 | 5 (62.5) | | | | |
| Apgar at five minutes (/10) | 9 (6.75, 9) | 9 (9, 9) | 9 (9, 9) | 0.25 | | 0.1 | |
| Apgar score <7 (n [%]) | 1 (25) | 0 | 0 | 0.24 | | 1 | |
| Missing (n [%]) | 0 | 0 | 5 (62.5) | | | | |
| **Placental anthropometry** | | | | | | | |
| Weight (g) | 495 (305, 633) [a] | 399 [ab] | 221 (147, 283) [b] | 0.03 | | 0.01 | |
| Missing (n [%]) | 0 | 7 (87.5) | 1 (12.5) | | | | |
| zWeight [*] | 1.46 (0.79, 2.88) | -2.35 | -0.62 (-1.24, 0.03) | 0.007 | | - | |
| Missing (n [%]) | 0 | 7 (87.5) | 2 (25) | | | | |
| Birthweight: placental weight ratio | 4.18 (3.23, 4.58) | 8.37 | 4.1 (3.53, 6.25) | 0.39 | | 0.03 | |
| Missing (n [%]) | 0 | 7 (87.5) | 1 (12.5) | | | | |
| **Placental pathology** | | | | | | | |
| % Intervillous fibrinoid | 1 (0, 4.25) | 5 (5, 12.5) | 5 (1.25, 8.75) | 0.06 | | 0.19 | |
| % Stellate reticulum | 27.5 (13.8, 41.3) | 10 (6.25, 26.3) | 40 (22.5, 67.5) | 0.05 | | 0.81 | |
| % Terminal villi | 30 (18.8, 30) | 32.5 (22.5, 38.8) | 15 (10, 36.3) | 0.39 | | 0.47 | |
| % Stem villi | 17.5 (15, 20) [a] | 10 (6.25, 10) [b] | 5 (5, 8.75) [b] | 0.004 | | 0.0002 | |
| % Mature intermediate villi | 12.5 (6.25, 15) | 10 (10, 10) | 7.5 (5, 10) | 0.13 | | 0.23 | |
| % Immature intermediate villi | 10 (6.25, 32.5) | 10 (5, 21.3) | 37.5 (15, 63.8) | 0.08 | | 0.15 | |
| Branching maturity score | 0.04 (0.01, 0.07) | 0.04 (0.02, 0.1) | 0.007 (0.002, 0.04) | 0.14 | | 0.81 | |
| Villous stromal maturity score | 0.09 (0.05, 0.23) | 0.18 (0.08, 0.44) | 0.05 (0.02, 0.09) | 0.04 | | 0.59 | |
| Syncytiotrophoblast maturity score | 2.5 (1.25, 3) | 4 (3, 4) | 1.5 (1, 3) | 0.03 | | 0.75 | |
| | | | | **Odds of placental pathology in cases (n=8)** | | | |
| | **Preterm controls (n=4)** | **SB controls (n=8)** | **Cases (n=8)** | **(vs. PT cons)** | | **(vs. SB cons)** | |
| **Placental pathology** | | | | **OR (95% CI)** | **aOR (95% CI)** [†] | **OR (95% CI)** | **aOR (95% CI)** [†] |
| Hofbauer cells (n [%]) | | | | | | | |
| Many | 1 (25) | 2 (25) | 6 (75) | 9 (0.7, 258) | 15.9 (0.21, 40640) | 9 (1.09, 115)* | 0.12 (0.0002, 6.57) |

| | | | | | | | |
|---|---|---|---|---|---|---|---|
| Few | 3 (75) | 6 (75) | 2 (25) | | | | |
| Cytotrophoblast density (n [%]) | | | | | | | |
| Thick | 1 (25) | 0 | 4 (50) | 3 (0.25, 76.6) | 0.79 (0.006, 46.9) | - | - |
| Thin/None | 3 (75) | 8 (100) | 4 (50) | | | | |
| Syncytiotrophoblast density (n [%]) | | | | | | | |
| Thick | 1 (25) | 1 (12.5) | 5 (62.5) | 5 (0.41, 131) | 5.23 (2.78, 1.44e8) | 11.7 (1.21, 283)* | 1.18e-5 (5.71e-17, 0.8) |
| Thin | 3 (75) | 7 (87.5) | 3 (37.5) | | | | |
| Vasculosyncytial membrane (n [%]) | | | | | | | |
| Many | 1 (25) | 3 (37.5) | 2 (25) | 1 (0.06, 26.8) | 8.07 (0.68, 28792) | 0.56 (0.05, 4.72) | 470 (2, 4.08e8) |
| Few/None | 3 (75) | 5 (62.5) | 6 (75) | | | | |
| Villi capillarisation (n [%]) | | | | | | | |
| Many | 0 | 2 (25) | 0 | - | - | - | - |
| Few/None | 4 (100) | 6 (75) | 8 (100) | | | | |
| Syncytial knots (n [%]) | | | | | | | |
| Many | 0 | 5 (63.5) | 1 (12.5) | - | - | 0.09 (0.004, 0.83)* | 1.5 (0.02, 165) |
| Few/None | 4 (100) | 3 (37.5) | 7 (87.5) | | | | |
| Collagen in stem villi (n [%]) | | | | | | | |
| Lots | 2 (50) | 3 (37.5) | 0 | - | - | - | - |
| Some/None | 2 (50) | 5 (62.5) | 8 (100) | | | | |
| Smooth muscle around stem vessels (n [%]) | | | | | | | |
| Lots | 2 (50) | 4 (50) | 1 (12.5) | 0.14 (0.005, 2.24) | 0.16 (0.002, 4.79) | 0.14 (0.006, 1.39) | 2.43 (0.05, 141) |
| Some/None | 2 (50) | 4 (50) | 7 (87.5) | | | | |
| Accelerated maturation (n [%]) | | | | | | | |
| Yes | 2 (50) | 0 | 1 (12.5) | 0.14 (0.005, 2.24) | 0.18 (0.003, 4.67) | - | - |
| No | 2 (50) | 8 (100) | 7 (87.5) | | | | |
| Distal villous hyperplasia (n [%]) | | | | | | | |
| Any | 2 (50) | 0 | 1 (12.5) | 0.14 (0.005, 2.24) | 0.18 (0.003, 4.67) | - | - |
| None | 2 (50) | 8 (100) | 7 (87.5) | | | | |
| Delayed villous maturation (n [%]) | | | | | | | |
| Yes | 0 | 2 (25) | 0 | - | - | - | - |
| No | 4 (100) | 6 (75) | 8 (100) | | | | |
| Maturation (n [%]) ‡ | | | | | | | |
| Immature | 0 | 3 (37.5) | 0 | - | - | - | - |
| Appropriate | 2 (50) | 5 (62.5) | 6 (75) | | | | |
| Hypermature | 2 (50) | 0 | 2 (25) | 0.33 (0.02, 4.31) | 0.46 (0.03, 7.3) | - | - |

† Adjusted for gestational age at delivery.
‡ Reference group: appropriate maturity.
Continuous data are median (IQR) with p value for one-way analysis of variance (Unadjusted: ANOVA [normal distribution/equal variance] or Wilcoxon test [non-parametric data or outliers]; Adjusted: Standard Least Squares models). Group differences from adjusted post-hoc tests (Tukey HSD) are indicated by different subscript letters. No letters = no group differences. Categorical data are n (%) and odds ratios or adjusted odds ratios (95% CI) for presence of pathology in case placentae (reference groups: PT controls and SB controls) from Nominal Logistic regression models with p value from Likelihood Ratio Chi Square test. A '-' indicates that data were unavailable. Statistical significance is denoted as *p<.05, **p<.01, ***p<.001. CI = confidence interval. PT = preterm. SB = spina bifida. TOP = termination of pregnancy. OR = odds ratio. aOR = adjusted odds ratio.
