## Supplementary Figures for "Altered placental phenotype and increased risk of placental pathology in fetal spina bifida: a matched case-control study"

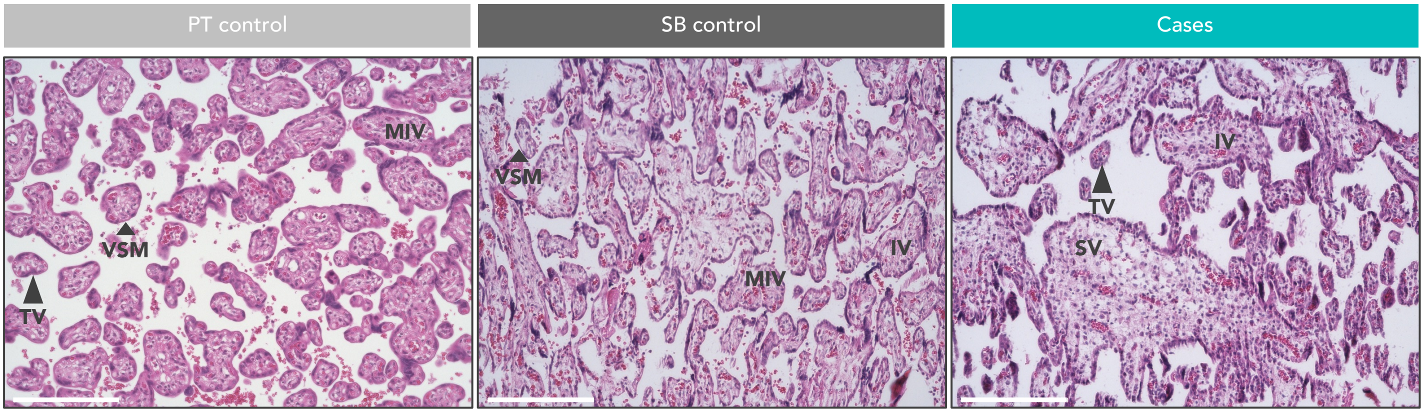


**Supplementary Figure S1. Representative images of H&E-stained placentae from PT controls, SB controls and cases.** IV = immature villus. TV = terminal villus (large arrowhead). SV = Stem villus. VSM = Vasculo-syncytial membrane (small arrowhead). MIV = mature intermediate villus. Scale bar = 200 μm. 20X magnification.

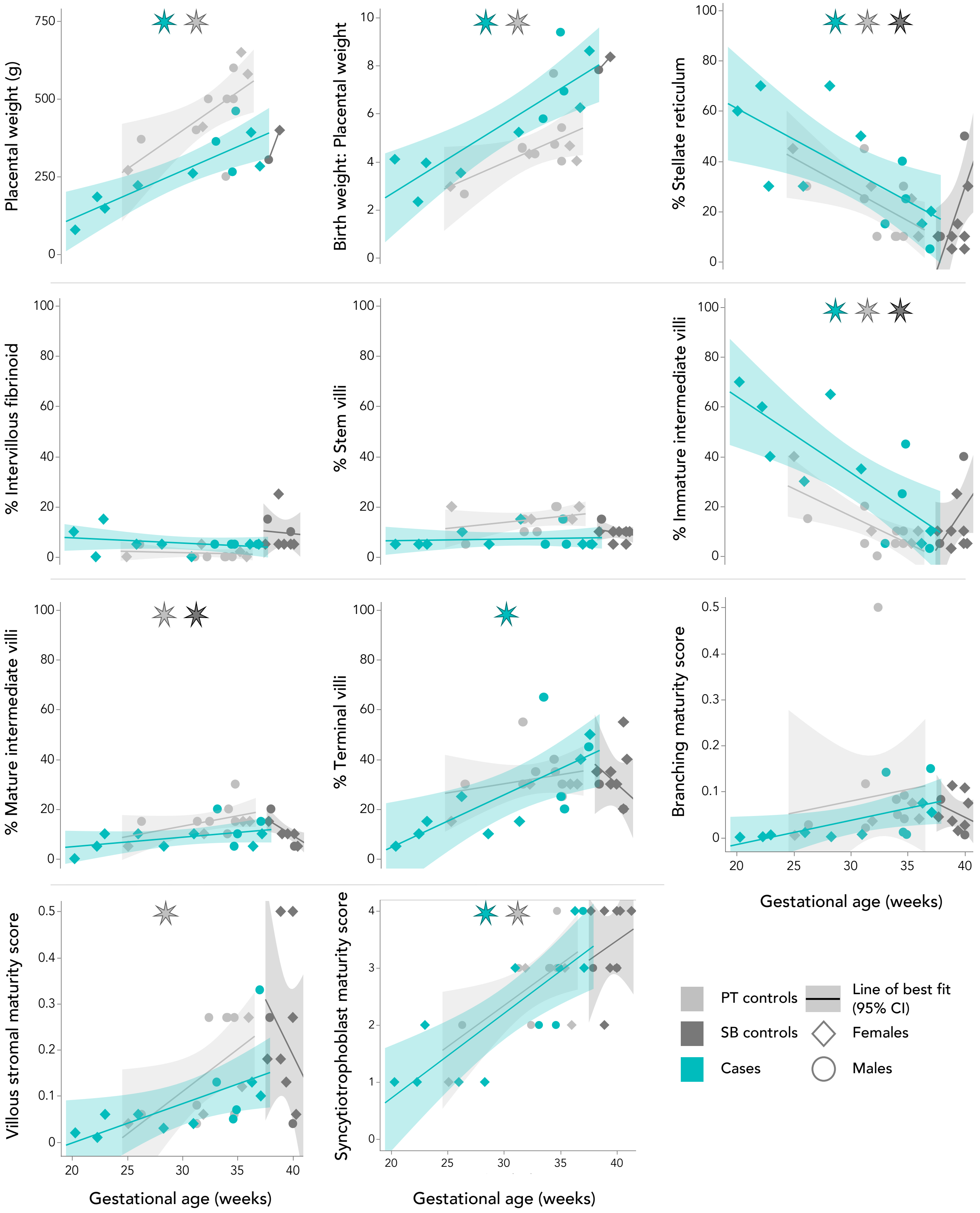


**Supplementary Figure S2. Associations between placental anthropometry, villi composition, overall maturity scores and gestational age at birth in PT controls, SB controls and cases.** Data are plotted by study group (light grey=PT controls, dark grey=SB controls, teal=cases) with line of best fit and 95% confidence interval from linear models. Significant associations from generalised linear models (normal distribution) are denoted by asterisks corresponding to study group colour ($p<0.05$). Diamonds = females. Circles = males.

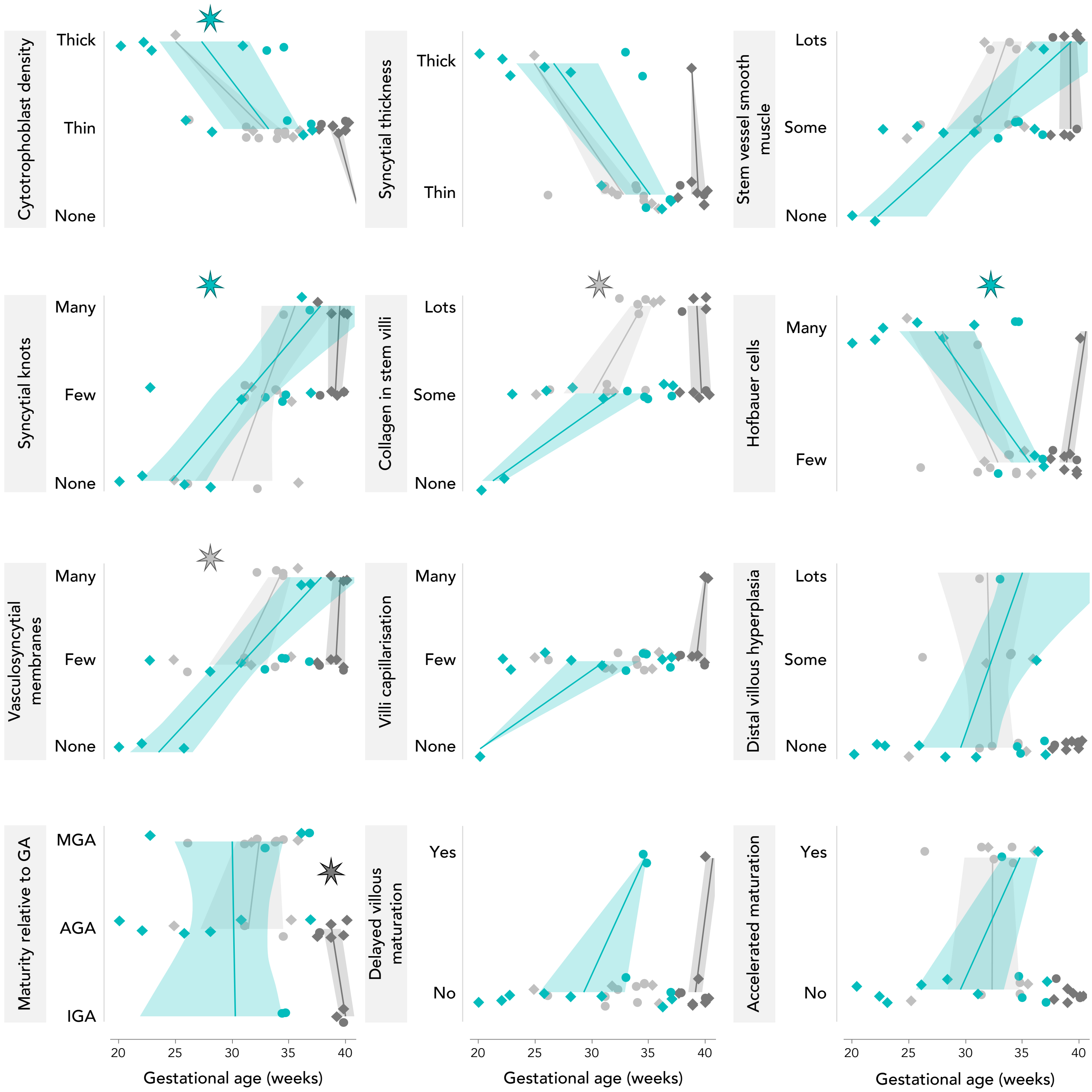


**Supplementary Figure S3. Associations between categorical placental histopathological characteristics and gestational age at birth in PT controls, SB controls and cases.** Data are plotted by study group (light grey=PT controls, dark grey=SB controls, teal=cases) with line of best fit and 95% confidence interval from linear models. Significant associations from generalised linear models (binomial distribution) are denoted by asterisks corresponding to study group colour (p<0.05). Diamonds = females. Circles = males.

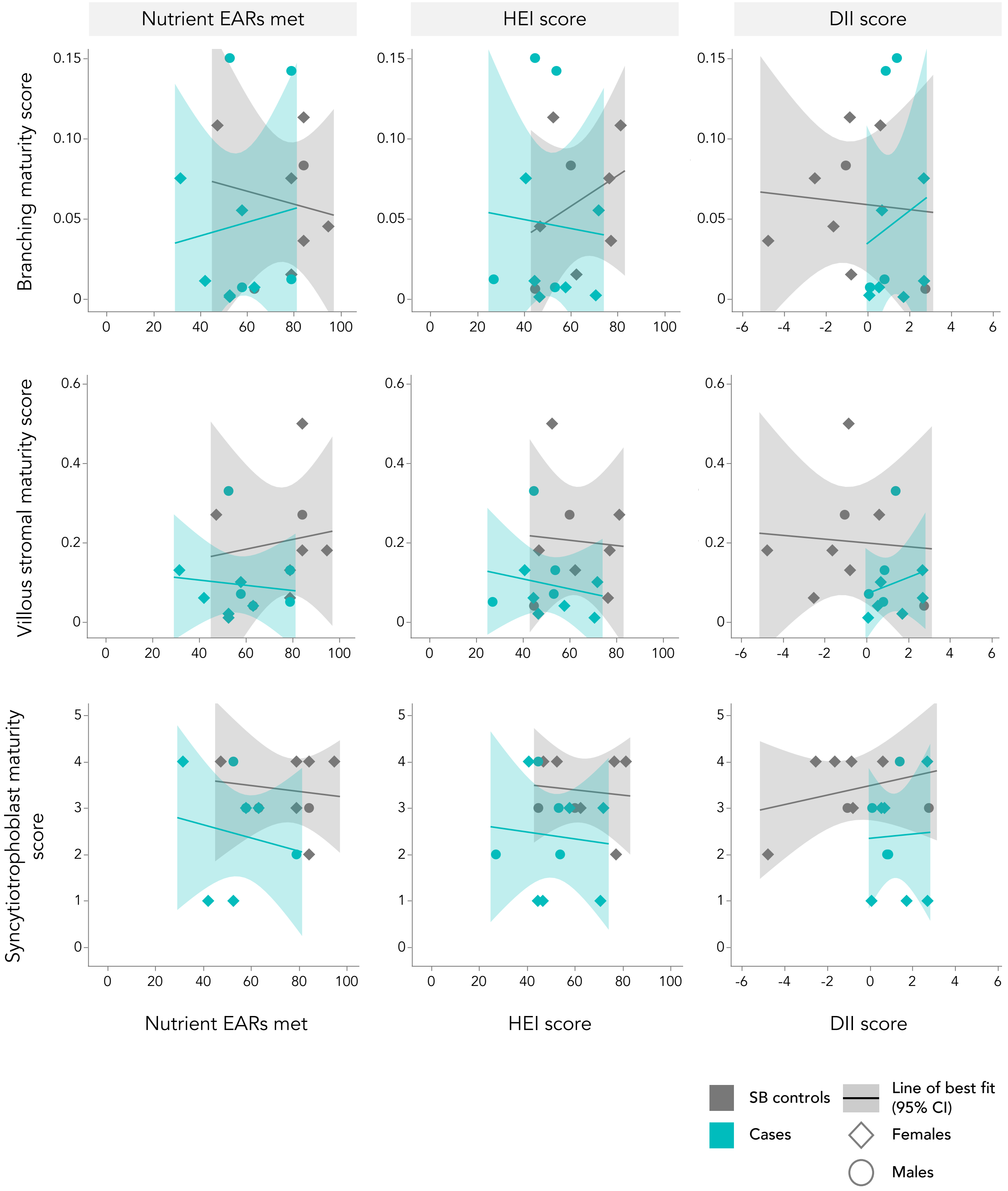


**Supplementary Figure S4. Summary measures of maternal dietary intakes in relation to placental maturity scores in cases and SB controls.** There were no associations between placental maturity scores and summary measures of maternal dietary intakes. EARs = estimated average requirements. HEI = health eating index. DII = dietary inflammatory index.
