## Supplementary Methods for "Altered placental phenotype and increased risk of placental pathology in fetal spina bifida: a matched case-control study"

### *Placental histopathology summary scores*

Summary scores for overall branching maturity, villous stromal maturity, stromal maturity and syncytiotrophoblast maturity were calculated (where a higher overall score indicates a more mature placenta, for all three measures). The branching maturity score considered the proportion of placental villi that were: terminal villi (%; higher proportion indicates higher maturity), mature intermediate villi (%; higher proportion indicates higher maturity), and immature intermediate villi (%; higher proportion indicates lower maturity). The villous stromal maturity score considered the amount of collagen in stem villi (none [scored as 0], some [1], a lot [2]; more collagen indicates higher maturity), the proportion of placental villi composed of stellate reticulum (%; higher proportion indicates lower maturity) and the level of Hofbauer cells in the villous stroma (few [scored as 1], many [2]; more Hofbauer cells indicates lower maturity). The syncytiotrophoblast maturity score considered the level of syncytial knots (none [scored as 0], few [1], many [2]; higher score indicates higher maturity) and the syncytial thickness (thick [scored as 1], thin [2]; higher score indicates higher maturity). The final equations were:

Eq. 1. Branching maturity score = ([terminal villi (%) + mature intermediate villi (%)] / [immature intermediate villi (%) + 1]) /100

Eq. 2. Villous stromal maturity score = (amount of collagen in stem villi + 1) / (stellate reticulum [%] + level of Hofbauer cells)

Eq. 3. Syncytiotrophoblast maturity score = level of syncytial knots + syncytial thickness

Summary scores for placental maturity were visualised as raw scores and transformed z-scores.

### *Transcriptome profiling*

To determine whether relationships exist between placental expression of gene pathways associated with NTDs and placental histopathology in cases, we evaluated relationships between placental histopathology and transcriptome signatures. The methods and results for microarray sequencing and gene-, pathway-, and network-level analyses of placental transcriptome in cases compared to PT and SB controls have been previously described (22). Here, these data were analysed in relation to placental histopathology.

First, to assess relationships between placental histopathology and overall (global) placental gene expression patterns, the top 20% of expressed, annotated transcripts from the Clariom™ D Human microarray were identified from previous analyses (n=17,251 transcripts; determined using the mean expression level after pre-processing across all samples) (22). Second, we used the list of genes that were differentially expressed in cases, compared to both PT and SB controls(22), to evaluate associations between placental histopathology and dysregulated gene expression in cases. Last, to assess relationships between placental histopathology and dysregulated gene pathway expression in cases, we determined associations between placental histopathology and normalised expression levels of genes associated with 14 previously identified geneset clusters of interest, that were up- or down- regulated in cases (22). The upregulated geneset clusters included pathways involved in angiogenesis and epithelial tube formation, extracellular matrix (ECM) activity, Hedgehog signaling, immune and inflammatory processes, protein synthesis, SH2 containing protein tyrosine phosphatase-2 (*SHP2*) activity, spinal cord injury, and Wnt signaling. The downregulated geneset clusters included pathways involved in Coat protein complex II (COPII)-mediated protein transport, endoplasmic reticulum (ER) activity, glycosylation, lipid metabolism, solute carrier (SLC) transport, and transmembrane transport. Data on expression levels of genes associated with each of these geneset clusters were used to evaluate associations between placental histopathology and dysregulated gene pathway activity in cases.

***Assessment of placental transcriptome features that discriminate placentae based on maturity***

In the first PCA approach, normalised expression levels of the top 20% of expressed, annotated transcripts were used to identify features of global placental gene expression that distinguished between placental samples. The top 10 principal components (PCs; 'global gene expression PCs'), of the total variation in transcriptome profile, were obtained and carried forward for further analysis. While the highest level of variance is typically explained by the first principal component (PC1), the top 10 PCs were considered because the variance explained by successive PCs, and the input variables (i.e., genes) most associated with these PCs, may also relate to outcomes of interest (e.g., measures of placental maturity). Standard least squares regression (for continuous maturity variables) and logistic regression (for categorical maturity variables) models, adjusted for infant sex and GA at delivery, were used to evaluate relationships between the top 10 global gene expression PCs and measures of placental maturity. Next, for each global gene expression PC that was significantly associated with measures of placental maturity ($p<0.05$), a list of genes with a loading value >0.4 (considered moderate/strong factor loadings (33, 35)) was obtained. To understand the functional profiles of global gene expression PCs that were significantly associated with measures of placental maturity, functional gene/protein- and pathway-level analysis was performed for genes with moderate/strong factor loadings using the STRING database. We focused on significantly enriched GO molecular functions, UniProt keywords and Reactome or Wikipathways (q value<0.05)(36).

In the second PCA approach, normalised expression levels of the DEGs between cases and all controls were used to determine how placental samples clustered based on the subset of genes that were differentially expressed in cases ('DEG PCA') (22). In the third PCA approach, PCA was performed using the normalised expression levels of the genes associated with each of the 14 geneset clusters of interest (i.e., 14 separate PCA models, each considering expression in a set of gene pathways with analogous function), to determine how placental samples clustered based on expression in differentially expressed gene pathways in cases (22) ('[geneset function] PCA'). From the DEG PCA model and each of the 14 geneset PCA models, only PC1 was obtained and carried forward for further analysis ('DEG PC1', '[geneset function] PC1'). As detailed functional analyses of the DEGs in cases has been previously reported (22), and the geneset-derived gene lists contained genes with similar and already know functions, only PC1, which accounted for the most variance in the data, was of interest. Standard least squares and logistic regression, adjusted for infant sex and GA at delivery, were used to evaluate relationships between measures of placental maturity and PCs representing the differentially expressed placental genes and gene pathways in cases.
